# Scaling the Prompt: How Batch Size Shapes Performance of Mid-2025 State-of-the-Art LLMs in Automated Title-and-Abstract Screening

**DOI:** 10.1101/2025.09.02.25334159

**Authors:** Petter Fagerberg, Oscar Sallander, Kim Vikhe Patil, Anders Berg, Anastasia Nyman, Natalia Borg, Thomas Lindén

**Affiliations:** The National Board of Health and Welfare; Institute of Neuroscience and Physiology, Sahlgrenska Academy, Gothenburg University, Sweden

**Keywords:** diagnostic test accuracy, large language models, artificial intelligence, meta-analysis, AI, systematic review, literature screening, Gemini, ChatGPT, validation

## Abstract

**Background:** Manual abstract screening is a primary bottleneck in evidence synthesis. Emerging evidence suggests that large language models (LLMs) can automate this task, but their performance when processing multiple records simultaneously in "batches" is uncertain.

**Objectives:** To evaluate the classification performance of four state-of-the-art LLMs (Gemini 2.5 Pro, Gemini 2.5 Flash, GPT-5, and GPT-5 mini) in predicting study eligibility across a wide range of batch sizes for a systematic review of randomised controlled trials.

**Methods:** We used a gold-standard dataset of 790 records (93 inclusions) from a published Cochrane Review. Using the public APIs for each model, batches of 1 to 790 citations were submitted to classify records as ’Include’ or ’Exclude’. Performance was assessed using sensitivity and specificity, with internal validation conducted through 10 repeated runs for each model-batch combination.

**Results:** Gemini 2.5 Pro was the most robust model, successfully processing the full 790-record batch. In contrast, GPT-5 failed at batches ≥400, while GPT-5 mini and Gemini 2.5 Flash failed at the 790-record batch. Overall, all models demonstrated strong performance within their operational ranges, with two notable exceptions: Gemini 2.5 Flash showed low initial sensitivity at batch 1, and GPT-5 mini’s sensitivity degraded at higher batch sizes (from 0.88 at batch 200 to 0.48 at batch 400). At a practical batch size of 100, Gemini 2.5 Pro achieved the highest sensitivity (1.00, 95% CI 1.00-1.00), whereas GPT-5 delivered the highest specificity (0.98, 95% CI 0.98-0.98).

**Conclusion:** State-of-the-art LLMs can effectively screen multiple abstracts per prompt, moving beyond inefficient single-record processing. However, performance is model-dependent, revealing trade-offs between sensitivity and specificity. Therefore, batch size optimisation and strategic model selection are important parameters for successful implementation.

## INTRODUCTION

Systematic reviews are the cornerstones of evidence-based medicine, but their production is a notoriously slow [1] and resource-intensive [2] process. The initial stage of title and abstract screening, a prognostic task which often involves manually sifting through thousands of records to predict their eligibility, represents a significant bottleneck [3]. The emergence of large language models (LLMs) offers a promising avenue for automating or augmenting this critical task [4], potentially accelerating evidence synthesis and reducing costs.

Prior research has demonstrated the potential of LLMs in systematic review screening [4], with some studies reporting high performance [5, 6]. However, these evaluations of existing models often share an important limitation: a reliance on a batch size of one, where each title and abstract is evaluated in a separate, individual application programming interface (API) call or chat prompt. This approach is inefficient for large-scale reviews and impractical for researchers without specialised programming skills, i.e. for those using the general user interface (GUI). Furthermore, the rapid pace of artificial intelligence (AI) development means that most published studies have evaluated LLMs that are now technologically outdated. Therefore, another justification for developing this new evaluation is the need to assess the performance of state-of-the-art (SOTA) models available in mid-2025.

The target population to use the prediction models evaluated in this study is researchers involved in evidence synthesis. The intended purpose of the current use of the LLMs is to automate the first-pass screening of titles and abstracts to support the decision of whether a study warrants full-text review. These tools are intended to be used at the beginning of the evidence synthesis pathway, following the initial literature search and deduplication, to triage citations into categories for inclusion, and exclusion, or further assessment.

The primary objective of this study was to evaluate the performance of four SOTA (mid-2025) LLMs, from two different AI model providers, for title-and-abstract screening. We aimed to quantify how classification performance changes as the number of citations processed in a single prompt (the "batch size") increases, thereby establishing the operational limits and optimal throughput for each model.

## METHODS

### Large Language Models

We evaluated two models each from two leading LLM families [7, 8]: Google and OpenAI [8–10]. All experiments were conducted between July 11 and August 8, 2025. The models were queried via their respective public APIs using default settings (i.e. for Gemini models temperature = 1.0, top_p = 1.0, and for OpenAI models reasoning effort = medium) to ensure a standardised and reproducible testing environment. Technical specifications for the models are provided in **Table 1**.

**Table 1.** Large language model specifications.

| Model Family | Model Name | API Identifier | Input Token Limit | Output Token Limit | Knowledge cutoff |
| --- | --- | --- | --- | --- | --- |
| Google | Gemini 2.5 Pro [9] | gemini-2.5-pro | 1,048,576 | 65,536 | January 2025 |
| Google | Gemini 2.5 Flash [10] | gemini-2.5-flash | 1,048,576 | 65,536 | January 2025 |
| OpenAI | GPT-5 (with thinking) [11] | gpt-5-2025-08-07 | 400,000 | 128,000 | October 2024 |
| OpenAI | GPT-5 mini (with thinking) [11] | gpt-5-mini-2025-08-07 | 400,000 | 128,000 | May 2024 |

### Data

This study utilised a single, purpose-built validation dataset derived from a systematic review in Cochrane Issue 6 (May-June 2025), "Stem cell treatment for acute myocardial infarction" [12]. To create an ecologically valid dataset simulating a real-world screening task, we supplemented the review’s publicly available EndNote library [12] by re-running the original search strategy. After deduplication, the final dataset comprised 790 records. This included 98 studies originally included in the review, 41 studies that were excluded only after full-text inspection, and 651 ’easy exclude’ records from the re-run search. The eligibility criteria for records were identical to those of the source Cochrane review, focusing on randomised controlled trials (RCTs) related to stem cell treatment for acute myocardial infarction. The validation dataset was chosen since it contained a relatively large number of included articles as well as being conducted according to Cochrane’s high standards (i.e. dual human abstract screeners).

### Experimental Workflow

To assess the impact of batch size on LLM performance, we submitted titles and abstracts in batches of 1, 10, 25, 50, 100, 150, 200, 400, and 790. We incrementally increased the batch size until the models failed to return a coherently formatted response, or the API request timed out. For each model and batch size combination, the screening process was repeated 10 times to establish stable performance estimates (mean, SD, 95% CIs).

The automated workflow proceeded as follows:

1. Reference data (study number, title, and abstract) were loaded from a source file.
2. A system prompt, the review-specific eligibility criteria, and a batch of references were programmatically combined.
3. The complete prompt was transmitted to the target LLM via its public API.
4. The API response was parsed. If the response did not conform to the expected structured table format (e.g., incorrect number of rows), the request was automatically re-attempted up to three times before being classified as a failure.

### LLM Prompting and Output Parsing

All models received the standardised system prompt shown in **Table 2**. A zero-shot prompting strategy was employed, meaning no examples of completed classifications were included in the prompt. This approach was chosen intentionally to isolate the effects of batch size rather than to optimise different prompting strategies. The prompt instructs the LLM to act as a systematic review expert, classify each study, and provide a one-sentence justification in a machine-readable markdown table.

**Table 2.** Standardised system prompt structure.

|  |
| --- |
| **System Prompt Sent to API:** |
| You are a systematic review expert. Does this study fulfil the criteria? Answer "Include" or "Exclude", if not sure answer "Full-text assessment" and give the reason why in one sentence max. If a study is a duplicate/from the same study as one included before, then decide "Include" again. Make your answers in a numbered table. Column 1 = the number of the study, column 2 = inclusion/exclusion/full text as one letter I E or F, column 3 = inclusion/exclusion reason, one sentence max. Make your answers in a numbered table.<br>Include study protocols, design/rationale papers and trial registry entries that describe an eligible RCT even if no results are yet reported; mark them as 'ongoing/awaiting classification'. Delivery modality (in-person, virtual, hybrid) is not an exclusion criterion unless explicitly stated. If any arm tests the intervention alone against a suitable control, include the study even if other arms contain additional co-interventions. Subgroup or secondary analyses based on an eligible RCT are eligible when they report outcomes for the target population. |
| **Criteria for considering studies for this review:** |
| "[insert systematic review criteria here]" |
| **List of titles/abstracts with study number:** |
| 1. Title 1 + Abstract 1 |
| 2. Title 2 + Abstract 2 |
| ... |
| 790. Title 790 + Abstract 790 |

### API Response Style

The prompt explicitly requested a markdown table to ensure a machine-readable format. Our parsing script was designed to handle this structure. If the API response did not conform to the expected tabular format (i.e., incorrect number of rows), the request was considered a failure and re- attempted. An example of a correctly formatted response from a model is shown in **Table 3**.

**Table 3.**
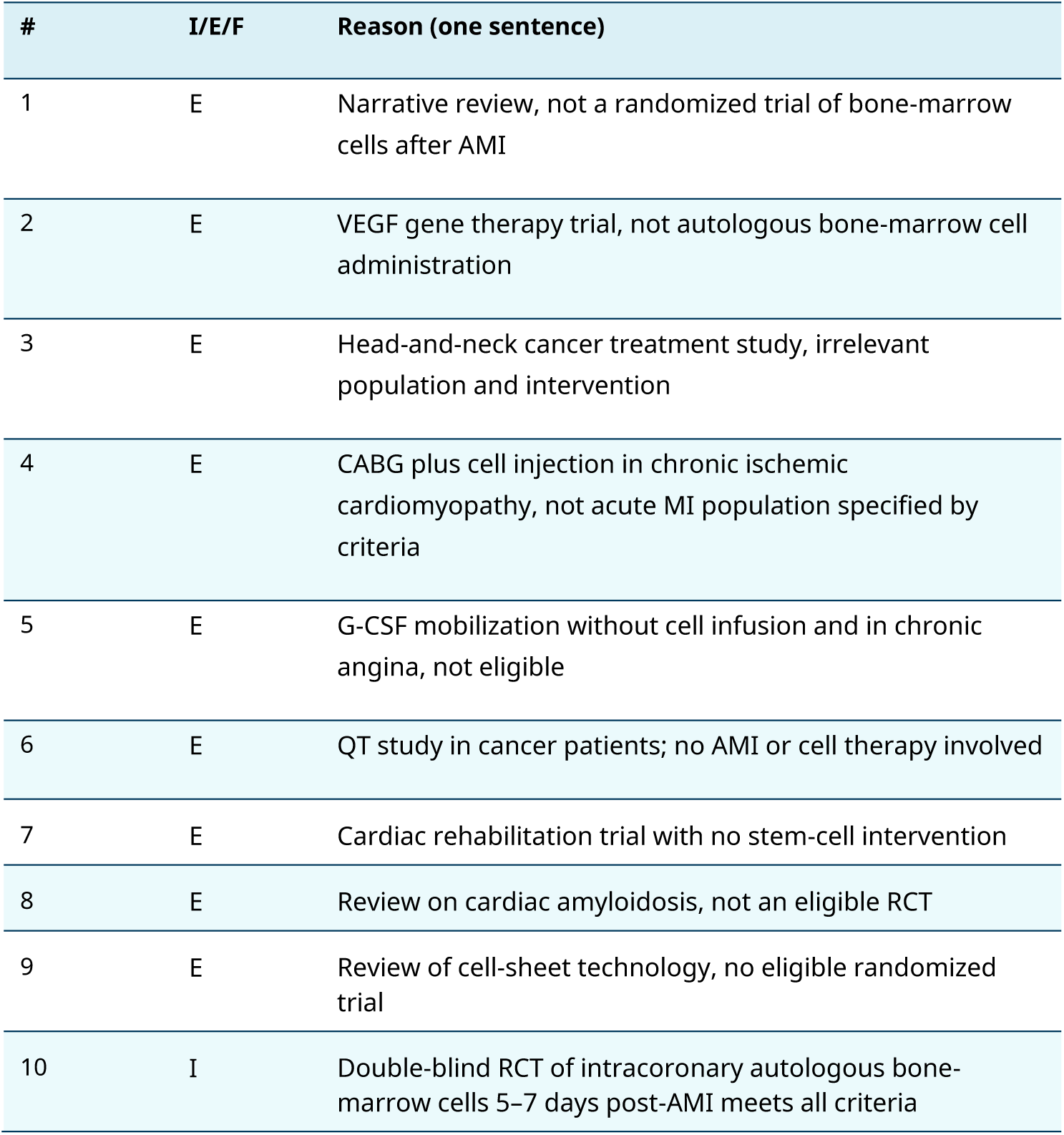
Illustrative example of a correct tabular output from an AI model (batch size 10).

### Outcome and Adjudication Process

We aimed to predict the eligibility of each record, classifying it as ’Include’ or ’Exclude’. While our initial gold standard was derived from the EndNote library of the source Cochrane review [12], we recognised that manual screening libraries can be imperfect. Therefore, we implemented a systematic, AI-assisted adjudication process to establish a more robust corrected gold standard. A disagreement was flagged when all 10 runs in batch size 1 of the state-of-the-art models (Gemini 2.5 Pro or GPT-5) unanimously contradicted the initial human-derived label. These flagged discrepancies were subsequently presented to two trained systematic reviewers, who considered the title and abstract, the original human decision, the unanimous AI decisions, and the study’s eligibility criteria to make a final determination (see appendix for detailed decisions). This adjudication process resulted in five records, originally labelled ’Include’, being re-classified as ’Exclude’. Consequently, our final validation dataset consisted of 93 included titles and abstracts, and all model performance metrics reported herein are benchmarked against this corrected gold standard.

### Statistical Analysis

Model performance was evaluated using standard classification metrics, defining true positives (TP) and true negatives (TN) as agreement with the corrected gold-standard, and false positives (FP) and false negatives (FN) as disagreement with the gold-standard. We calculated:

- Sensitivity = TP / (TP + FN)
- Specificity = TN / (TN + FP)

For each metric, we reported the mean, standard deviation (SD), and standard error (SE) across the 10 independent runs (see appendix for detailed results). The 95% confidence intervals (CIs) for the mean of each metric were calculated using a t-distribution. All statistical calculations and data visualizations were performed using R (version 4.5.1), with key packages including dplyr for data manipulation and ggplot2 for plotting. [14].

## RESULTS

### Records

The study flow involved screening records from a re-executed literature search combined with an existing review library. After deduplication, a final set of 790 unique records was included in the analysis. Of these, 93 records were true inclusions (positive outcome events) in the corrected gold standard, resulting in an outcome prevalence of 11.8%. The remaining 697 records were true exclusions.

### Model Performance

#### Model performance was evaluated across nine batch sizes

(1, 10, 25, 50, 100, 150, 200, 400, and 790 titles/abstracts per prompt), with 10 replicates per condition, leaving 7900 unique decisions for each batch size for each AI model.

#### Response Validity and Maximum Workable Batch Size

All four models produced syntactically valid, machine-readable tables for batch sizes up to and including 200 abstracts. Gemini 2.5 Pro was the only model that remained fully reliable across the entire range, successfully processing the full-corpus batch of 790 abstracts in all 10 runs. Gemini 2.5 Flash and GPT-5 mini maintained 100% response validity up to 400 abstracts but failed at 790. GPT-5 showed instability at higher loads, failing completely at 400 (even after multiple retries).

### Sensitivity

Gemini 2.5 Pro was the most sensitive model, maintaining a mean sensitivity ≥0.99 for batch sizes below 790 and achieving 0.98 (95% CI 0.97-0.99) even at the full-corpus batch size of 790 (**Figure 1**). Similarly, GPT-5 exhibited stable sensitivity (range of means 0.97-0.99) at all batch sizes, except for an initial value of 0.95 (95% CI 0.94-0.96) at batch 1. In contrast, the other models showed more variable performance. GPT-5 mini’s sensitivity dropped sharply from 0.88 (95% CI 0.81-0.95) at batch 200 to 0.48 (95% CI 0.39-0.57) at batch 400, indicating its effective context limit was reached at approximately batch 150. Gemini 2.5 Flash also showed reduced sensitivity at batch 1 (0.85; 95% CI 0.85-0.86) but improved with larger batches, converging with GPT-5’s performance by batch 200 (0.98; 95% CI 0.96-0.99).

**Figure 1.**
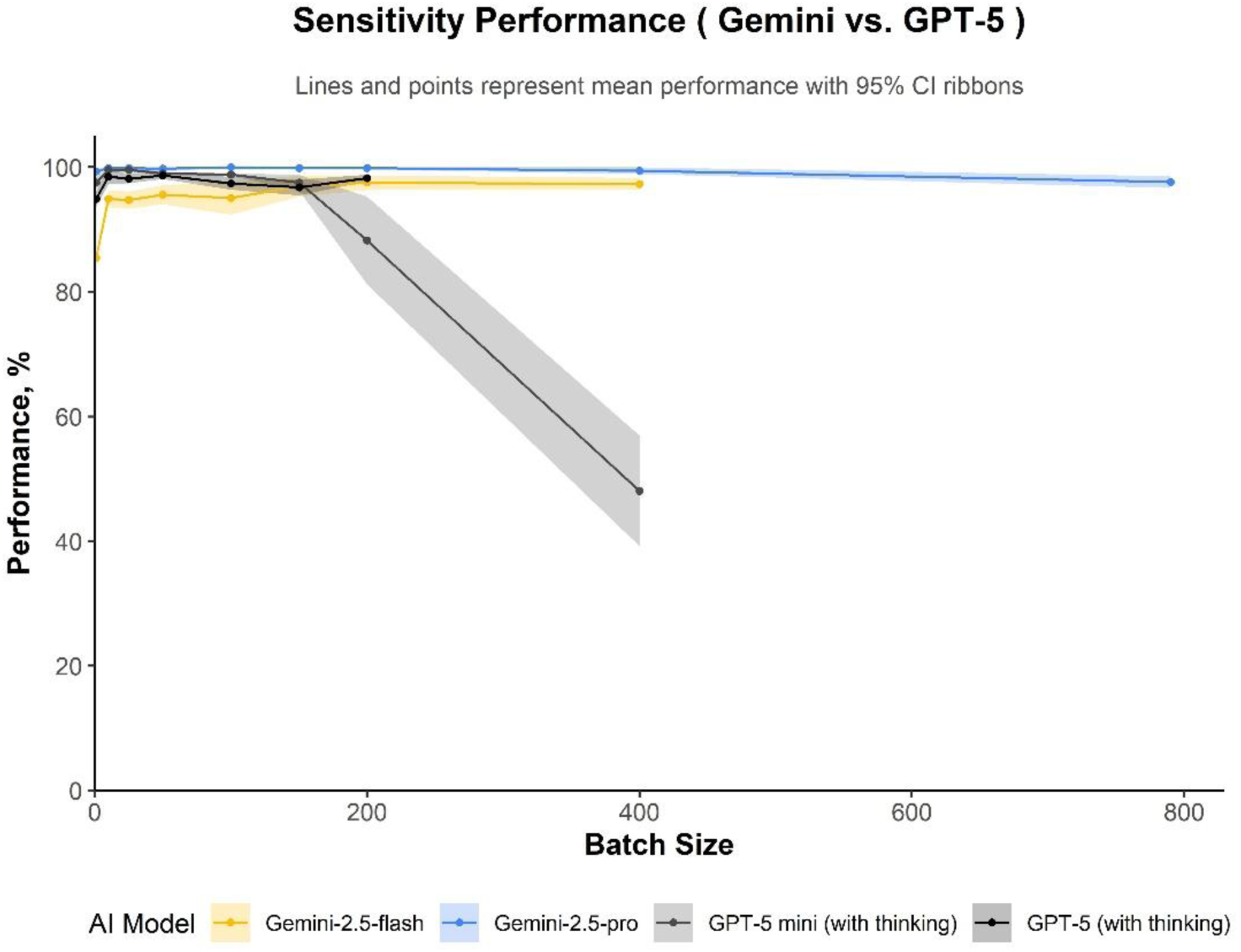
Sensitivity across batch sizes (95 % CIs), n = 7900 decision at each plotted batch size for each AI model. Mean sensitivity (points) with 95 % CIs (shaded). True-positive rates for each model.

### Specificity

GPT-5 was the most specific model across all batch sizes (range 0.98-0.99, **Figure 2**). GPT-5 mini also maintained high specificity (range 0.96-0.98), with a notable exception at batch 400 where it dropped to 0.92 (95% CI 0.81-1.00). In contrast, the Gemini models exhibited more variable performance that was dependent on batch size. After batch 1, Gemini 2.5 Pro’s specificity peaked at a batch size of 150 (0.96; 95% CI 0.95-0.96), while Gemini 2.5 Flash reached its maximum specificity much earlier at batch 10 (0.94; 95% CI 0.94-0.95).

**Figure 2.**
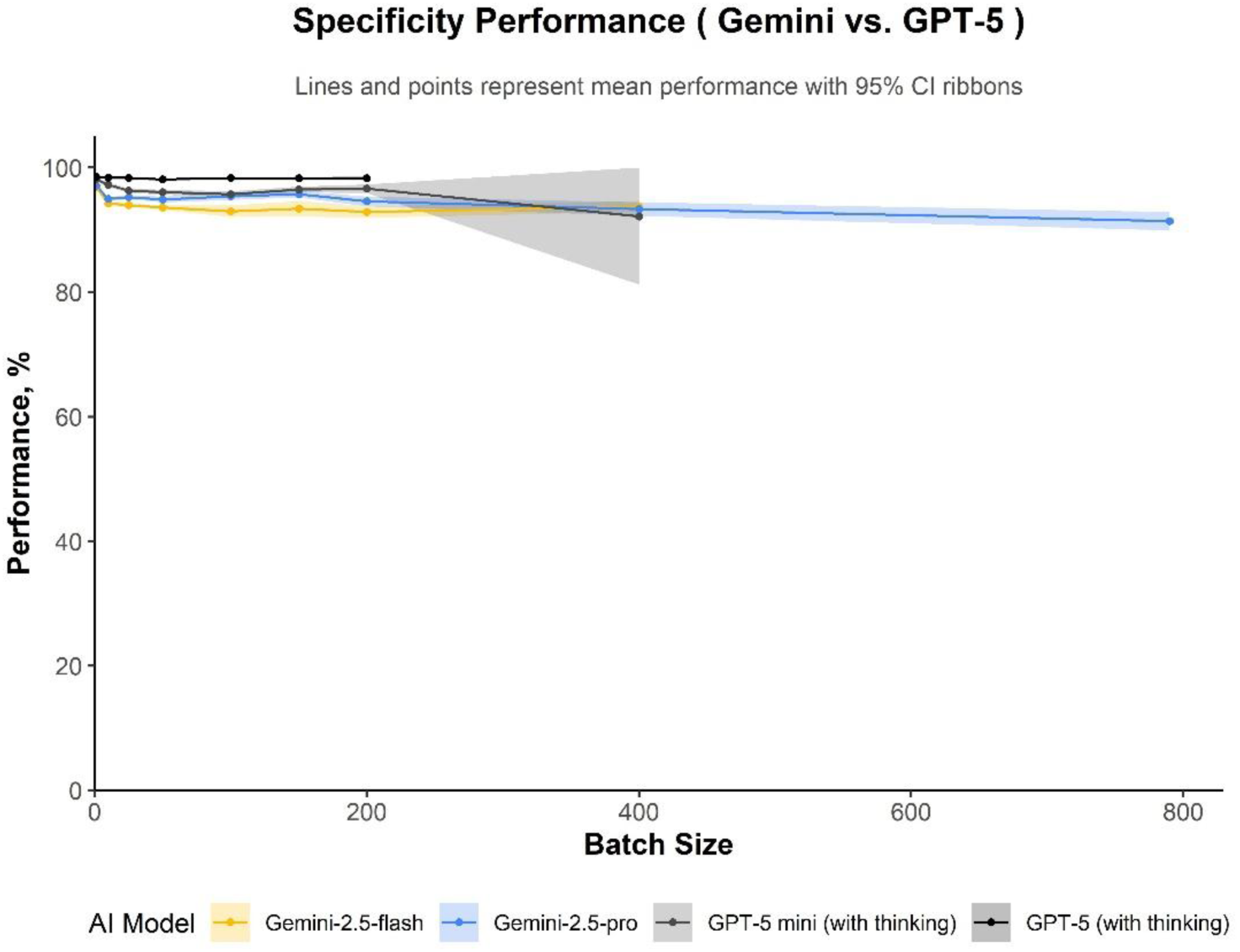
Specificity across batch sizes (95 % CIs), n = 7900 decision at each plotted batch size for each AI model. Mean sensitivity (points) with 95 % CIs (shaded). True-negative rates for each model.

Overall, the AI models tested in this study demonstrated robust performance across most of tested batch sizes. The two notable exceptions to this trend were GPT-5 mini, which showed a significant performance decline at batch size 200 and 400, and Gemini 2.5 Flash, which exhibited reduced sensitivity specifically at batch size 1 (see detailed results in appendix).

For a practical application, such as a non-technical user operating via a general user interface with a batch size of 100, Gemini 2.5 Pro offered the highest sensitivity (1.00 95% CI 1.00-1.00), while GPT-5 achieved the highest specificity (0.98 95% CI 0.98-0.98), as detailed in **Table 4**.

**Table 4.**
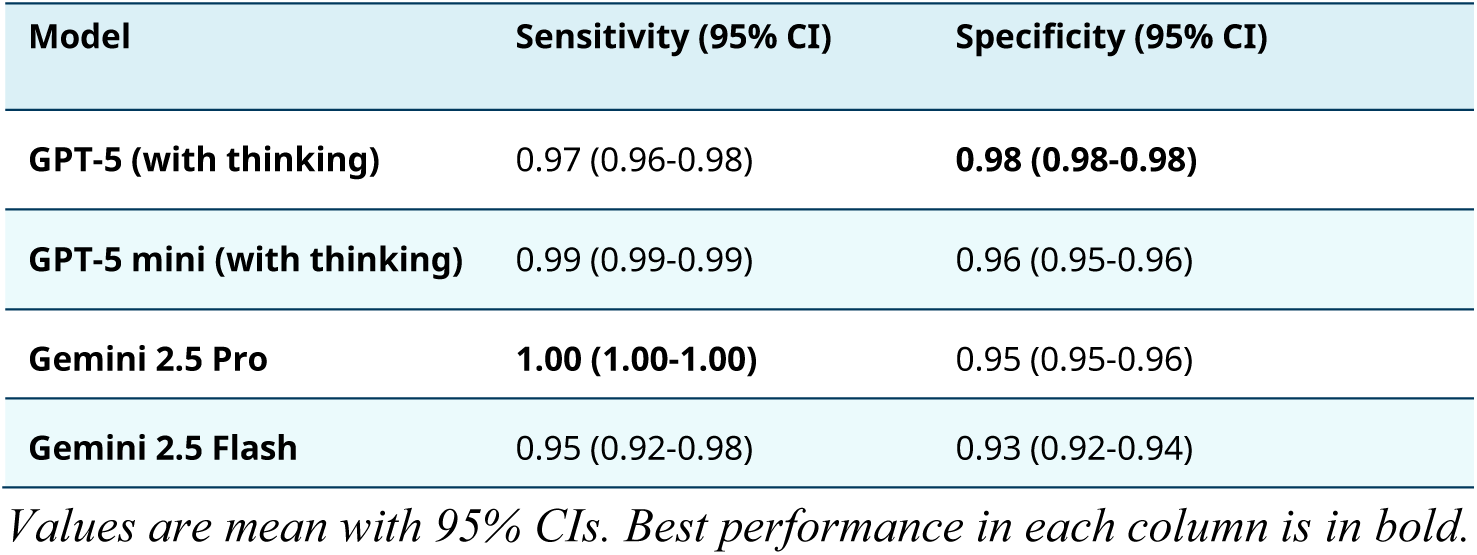
Model performance at a matched batch size of 100 titles and abstracts per prompt. A practical batch size for a non-technical user in the general user interface.

## DISCUSSION

This study is the first to demonstrate that contemporary (mid 2025) large language models (LLMs) can accurately screen titles and abstracts in large batches in the systematic review process, representing a methodological advance over more inefficient single-record processing. By systematically evaluating performance across various batch sizes; an important operational parameter largely unaddressed in previous reports [4], we show that high-throughput, batch-based screening is viable without a substantive loss of performance. However, our findings reveal distinct, model-dependent performance profiles that can inform model selection. Gemini 2.5 Pro proved highly robust across the widest range of batch sizes, indicating its suitability for high-volume screening where high sensitivity is crucial.

Conversely, GPT-5 and GPT-5 mini offered superior specificity, making them preferable for workflows where minimizing false positives is paramount. This heterogeneity provides an evidence-based rationale for tailoring the choice of LLM to specific review priorities and suggests the potential for using model ensembles to mitigate idiosyncratic biases and error patterns.

This study has several limitations that warrant consideration. First, our findings are based on a single, topic-specific dataset of randomised controlled trials for acute myocardial infarction. This reliance on one clinical domain and study type may limit the generalisability of our performance estimates to other areas of medical research. Second, while we selected a recent systematic review as a validation dataset to minimise data contamination, a potential for temporal overlap with the models’ training data remains. The source review was published online in 2024 [12], with several earlier versions being published previously as well, and because the exact knowledge cut-off dates for the mid-2025 models are after this time of publication, we cannot definitively exclude the possibility that performance was influenced by prior exposure to the review’s inclusion decisions.

However, it is important to contextualise this limitation. As our primary objective was to evaluate the relative effects of batch size, this potential contamination is of less concern; the performance trends observed are likely independent of prior data exposure, since it is the same AI models being tested across different batch sizes. For future studies aiming to validate the absolute performance of these models for real-world use, employing datasets published unequivocally after the models’ training dates will be important.

Third, our study deliberately employed a standardised, zero-shot prompt to isolate the effect of batch size. The reported accuracies may not represent the models’ maximum capabilities, as performance could likely be enhanced using more optimised few-shot, multi-shot (providing the model with examples of difficult-to-classify cases) or other more advanced prompting strategies [5]. An investigation into such prompt engineering, however, was beyond the scope of this research. Fourth, our AI-assisted adjudication process resulted in the re-classification of five records from ’Include’ to ’Exclude’. This discrepancy does not appear to represent screening errors in the source review. Rather, it is likely a "carry-over effect" from the original authors’ transparent practice of including all associated publications (i.e., protocols for eligible studies, sub-studies etc.) in their EndNote library, even if these specific records were not used in their final quantitative synthesis.

This finding underscores the models’ ability to adhere strictly to the given eligibility criteria, ignoring the contextual associations that might lead a human to retain such supplementary files. Creating this ‘corrected’ gold standard was essential for our study, as our primary goal was to evaluate AI performance against a precise eligibility criterion rather than to replicate the original, more comprehensive EndNote library. This AI-assisted adjudication, therefore, ensured a more robust and valid benchmark for our performance metrics. Fifth, our analysis excluded runs that failed to produce a syntactically valid output after three attempts. This approach may slightly inflate the reported accuracy for some models at their operational limits by not penalising them for catastrophic failures. For example, GPT-5 mini needed one retry before being able to complete all 10 runs at batch 400.

Finally, the rapid evolution of commercial large language models presents a significant challenge to the long-term reproducibility of our findings. For instance, during our study period (July-August 2025), the models initially tested (i.e., o3 and GPT-4.1) were superseded and deprecated (in the general user interface) by the provider in favour of a new model family (GPT-5).

This rapid iteration underscores that performance metrics are specific to a model version at a particular point in time and may not generalise to future releases (see Appendix for a performance of all tested AI models). To address this challenge, several strategies can be employed. One approach is to use stable, version-controlled models. Open-source models that can be run locally are particularly advantageous, as they insulate research from provider-driven updates and depreciation schedules. For studies that rely on commercial models, a complementary strategy is the regular use of a standardised benchmark dataset, like the one developed here, to monitor for performance drift from unannounced "silent" updates. Furthermore, researchers using these commercial services might consider committing to stable API versions and only migrate to new releases after conducting thorough re-validation.

Translating these findings into practice requires consideration of user expertise, workflow integration, and future development. For practical implementation, users must handle data imperfections, as records with missing or poor-quality abstracts will require separate manual review. Although the current methodology demands technical skill in handling APIs and scripting, domain knowledge is also critical. Reviewers with subject matter expertise can better tailor the eligibility criteria and interpret the AI’s rationales, creating a feedback loop to iteratively refine screening protocols. This highlights a crucial point: the LLM’s performance is highly dependent on the detail and clarity of the initial review criteria, meaning greater emphasis should be dedicated to this foundational step of the process.

Based on our findings, a human-in-the-loop implementation appears to be a promising direction for future work. Research could explore a workflow where ensemble LLMs performs a first-pass screening using optimised batch sizes, such as 100 abstracts for Gemini 2.5 Pro and GPT-5. A key aspect to investigate would be the impact of subsequent human verification of a sample (i.e., 5-10%) of AI-generated exclusions. Such a hybrid system presents a testable model for balancing the efficiency of automation with expert supervision.

Looking forward, a crucial next step involves evaluating the practical usability of existing graphical user interfaces (GUIs) for this batch- prompting workflow. Future research should investigate how easily the average systematic reviewer can learn and effectively apply these tools for large-batch screening, as this is key to lowering the current technical barrier and promoting wider adoption. Beyond usability testing, the models are ready for evaluation in prospective AI-assisted systematic reviews. Such studies should be designed to formally compare this AI-assisted workflow against the traditional manual screening process, measuring key differences in cost-effectiveness, time saved, and overall screening accuracy.

Furthermore, the rapid pace of AI development suggests the batch-processing workflow evaluated in this paper represents a transient methodology. As of mid-2025, this development is characterised by two synergistic advancements: qualitative improvements in core reasoning capabilities and the expansion of context windows. More advanced versions of the tested models (although more expensive to use), like Gemini 2.5 Deep Think [15] and GPT-5 Pro [16] for instance, demonstrate enhanced analytical capabilities through novel computational approaches, a trend expected to continue with forthcoming models such as Gemini 3.0 and GPT-6. Concurrently, the expansion of context windows; projected to reach two million tokens for models like Gemini 2.5 Pro soon [17], creates the technical possibility for processing substantially larger inputs. The convergence of these trends raises the possibility of corpus-level abstract screening, where an entire bibliographic database could theoretically be analysed within a single prompt. The viability of such an approach, however, depends on critical methodological challenges. Notably, these include the "needle-in-a-haystack" problem [18] and “context rot” [19], which describe the difficulty of maintaining reliable information retrieval across vast token lengths. Consequently, while this study validates an effective workflow for contemporary models mid-2025, it also functions as a benchmark. It establishes a performance baseline against which the feasibility of future, more integrated screening methods can be assessed as the underlying technology continues to mature.

## CONCLUSION

Our findings demonstrate that state-of-the-art large language models (as of mid-2025) can effectively screen multiple records per prompt in systematic reviews. High performance, however, is dependent upon optimising the batch size to each model’s specific context tolerance. The performance profile is model-dependent: Gemini 2.5 Pro offers versatile, high-throughput sensitivity capabilities, while GPT-5 model family excels in specificity. This contrast points to model ensembles as a promising strategy for balancing competing performance objectives and reducing idiosyncratic model error risks. To ensure the generalisability of these findings, the critical next step is to validate these AI-assisted workflows across a wider range of diverse datasets.

## Data Availability

All data produced in the present study are available upon reasonable request to the authors

## Acknowledgements

We thank the authors of the original Cochrane Review for their rigorous work and for making their EndNote libraries available.

## Ethics approval and consent to participate

Not applicable.

## Author contributions

PF and OS had full access to all the data in the study and took responsibility for the integrity of the data and the data analysis.

Concept and design: PF, OS and KVP. Acquisition, analysis, or interpretation of data: PF and OS. Drafting of the manuscript: PF, KVP and OS. Critical review of the manuscript for important intellectual content: All authors. Statistical analysis: PF and OS. Administrative, technical, or material support: AB, AN, TL, NB. Supervision: AB, AN, TL, NB. All authors read and approved the definitive version of the manuscript.

## Competing interests

None.

## Data availability statement

The datasets used and analysed in this study are available upon reasonable request.

## Funding statement

The authors received no specific funding for this work.

## APPENDIX

### Sensitivity

| AI Model | Batch Size | Mean | Std Dev | Std Error | Lower 95% CI | Upper 95% CI |
| --- | --- | --- | --- | --- | --- | --- |
| Gemini-2.5-flash | 1 | 0.85 | 0.01 | 0.00 | 0.85 | 0.86 |
| Gemini-2.5-flash | 10 | 0.95 | 0.02 | 0.01 | 0.94 | 0.96 |
| Gemini-2.5-flash | 25 | 0.95 | 0.02 | 0.01 | 0.93 | 0.96 |
| Gemini-2.5-flash | 50 | 0.96 | 0.02 | 0.01 | 0.94 | 0.97 |
| Gemini-2.5-flash | 100 | 0.95 | 0.04 | 0.01 | 0.92 | 0.98 |
| Gemini-2.5-flash | 150 | 0.97 | 0.02 | 0.01 | 0.95 | 0.99 |
| Gemini-2.5-flash | 200 | 0.98 | 0.02 | 0.01 | 0.96 | 0.99 |
| Gemini-2.5-flash | 400 | 0.97 | 0.01 | 0.00 | 0.96 | 0.98 |
| Gemini-2.5-pro | 1 | 0.99 | 0.01 | 0.00 | 0.99 | 1.00 |
| Gemini-2.5-pro | 10 | 1.00 | 0.00 | 0.00 | 1.00 | 1.00 |
| Gemini-2.5-pro | 25 | 1.00 | 0.00 | 0.00 | 1.00 | 1.00 |
| Gemini-2.5-pro | 50 | 1.00 | 0.01 | 0.00 | 0.99 | 1.00 |
| Gemini-2.5-pro | 100 | 1.00 | 0.00 | 0.00 | 1.00 | 1.00 |
| Gemini-2.5-pro | 150 | 1.00 | 0.00 | 0.00 | 1.00 | 1.00 |
| Gemini-2.5-pro | 200 | 1.00 | 0.00 | 0.00 | 1.00 | 1.00 |
| Gemini-2.5-pro | 400 | 0.99 | 0.01 | 0.00 | 0.99 | 1.00 |
| Gemini-2.5-pro | 790 | 0.98 | 0.01 | 0.00 | 0.97 | 0.99 |
| GPT-5 mini (with thinking) | 1 | 0.98 | 0.01 | 0.00 | 0.97 | 0.98 |
| GPT-5 mini (with thinking) | 10 | 1.00 | 0.01 | 0.00 | 0.99 | 1.00 |
| GPT-5 mini (with thinking) | 25 | 1.00 | 0.01 | 0.00 | 0.99 | 1.00 |
| GPT-5 mini (with thinking) | 50 | 0.99 | 0.00 | 0.00 | 0.99 | 0.99 |
| GPT-5 mini (with thinking) | 100 | 0.99 | 0.00 | 0.00 | 0.99 | 0.99 |
| GPT-5 mini (with thinking) | 150 | 0.98 | 0.02 | 0.01 | 0.96 | 0.99 |
| GPT-5 mini (with thinking) | 200 | 0.88 | 0.10 | 0.03 | 0.81 | 0.95 |
| GPT-5 mini (with thinking) | 400 | 0.48 | 0.12 | 0.04 | 0.39 | 0.57 |
| GPT-5 (with thinking) | 1 | 0.95 | 0.02 | 0.01 | 0.94 | 0.96 |
| GPT-5 (with thinking) | 10 | 0.98 | 0.02 | 0.01 | 0.97 | 1.00 |
| GPT-5 (with thinking) | 25 | 0.98 | 0.01 | 0.00 | 0.97 | 0.99 |
| GPT-5 (with thinking) | 50 | 0.99 | 0.01 | 0.00 | 0.98 | 0.99 |
| GPT-5 (with thinking) | 100 | 0.97 | 0.01 | 0.00 | 0.96 | 0.98 |
| GPT-5 (with thinking) | 150 | 0.97 | 0.02 | 0.01 | 0.96 | 0.98 |
| GPT-5 (with thinking) | 200 | 0.98 | 0.01 | 0.00 | 0.98 | 0.99 |

### Specificity

| AI Model | Batch Size | Mean | Std Dev | Std Error | Lower 95% CI | Upper 95% CI |
| --- | --- | --- | --- | --- | --- | --- |
| Gemini-2.5-flash | 1 | 0.97 | 0.00 | 0.00 | 0.97 | 0.97 |
| Gemini-2.5-flash | 10 | 0.94 | 0.01 | 0.00 | 0.94 | 0.95 |
| Gemini-2.5-flash | 25 | 0.94 | 0.00 | 0.00 | 0.94 | 0.94 |
| Gemini-2.5-flash | 50 | 0.94 | 0.00 | 0.00 | 0.93 | 0.94 |
| Gemini-2.5-flash | 100 | 0.93 | 0.01 | 0.00 | 0.92 | 0.94 |
| Gemini-2.5-flash | 150 | 0.93 | 0.02 | 0.01 | 0.92 | 0.95 |
| Gemini-2.5-flash | 200 | 0.93 | 0.01 | 0.00 | 0.92 | 0.94 |
| Gemini-2.5-flash | 400 | 0.94 | 0.01 | 0.00 | 0.93 | 0.95 |
| Gemini-2.5-pro | 1 | 0.97 | 0.00 | 0.00 | 0.97 | 0.97 |
| Gemini-2.5-pro | 10 | 0.95 | 0.00 | 0.00 | 0.95 | 0.95 |
| Gemini-2.5-pro | 25 | 0.95 | 0.00 | 0.00 | 0.95 | 0.96 |
| Gemini-2.5-pro | 50 | 0.95 | 0.01 | 0.00 | 0.94 | 0.95 |
| Gemini-2.5-pro | 100 | 0.95 | 0.01 | 0.00 | 0.95 | 0.96 |
| Gemini-2.5-pro | 150 | 0.96 | 0.01 | 0.00 | 0.95 | 0.96 |
| Gemini-2.5-pro | 200 | 0.95 | 0.01 | 0.00 | 0.94 | 0.95 |
| Gemini-2.5-pro | 400 | 0.93 | 0.02 | 0.00 | 0.92 | 0.94 |
| Gemini-2.5-pro | 790 | 0.91 | 0.02 | 0.01 | 0.90 | 0.93 |
| GPT-5 mini (with thinking) | 1 | 0.98 | 0.00 | 0.00 | 0.98 | 0.98 |
| GPT-5 mini (with thinking) | 10 | 0.97 | 0.01 | 0.00 | 0.97 | 0.98 |
| GPT-5 mini (with thinking) | 25 | 0.96 | 0.00 | 0.00 | 0.96 | 0.97 |
| GPT-5 mini (with thinking) | 50 | 0.96 | 0.01 | 0.00 | 0.96 | 0.96 |
| GPT-5 mini (with thinking) | 100 | 0.96 | 0.01 | 0.00 | 0.95 | 0.96 |
| GPT-5 mini (with thinking) | 150 | 0.96 | 0.01 | 0.00 | 0.96 | 0.97 |
| GPT-5 mini (with thinking) | 200 | 0.97 | 0.01 | 0.00 | 0.96 | 0.97 |
| GPT-5 mini (with thinking) | 400 | 0.92 | 0.15 | 0.05 | 0.81 | 1.00 |
| GPT-5 (with thinking) | 1 | 0.99 | 0.00 | 0.00 | 0.98 | 0.99 |
| GPT-5 (with thinking) | 10 | 0.98 | 0.00 | 0.00 | 0.98 | 0.99 |
| GPT-5 (with thinking) | 25 | 0.98 | 0.00 | 0.00 | 0.98 | 0.98 |
| GPT-5 (with thinking) | 50 | 0.98 | 0.00 | 0.00 | 0.98 | 0.98 |
| GPT-5 (with thinking) | 100 | 0.98 | 0.00 | 0.00 | 0.98 | 0.98 |
| GPT-5 (with thinking) | 150 | 0.98 | 0.00 | 0.00 | 0.98 | 0.98 |
| GPT-5 (with thinking) | 200 | 0.98 | 0.00 | 0.00 | 0.98 | 0.99 |

### Accuracy

| AI Model | Batch Size | Mean | Std Dev | Std Error | Lower 95% CI | Upper 95% CI |
| --- | --- | --- | --- | --- | --- | --- |
| Gemini-2.5-flash | 1 | 0.96 | 0.00 | 0.00 | 0.95 | 0.96 |
| Gemini-2.5-flash | 10 | 0.94 | 0.01 | 0.00 | 0.94 | 0.95 |
| Gemini-2.5-flash | 25 | 0.94 | 0.01 | 0.00 | 0.94 | 0.94 |
| Gemini-2.5-flash | 50 | 0.94 | 0.00 | 0.00 | 0.94 | 0.94 |
| Gemini-2.5-flash | 100 | 0.93 | 0.01 | 0.00 | 0.92 | 0.94 |
| Gemini-2.5-flash | 150 | 0.94 | 0.02 | 0.01 | 0.93 | 0.95 |
| Gemini-2.5-flash | 200 | 0.93 | 0.01 | 0.00 | 0.93 | 0.94 |
| Gemini-2.5-flash | 400 | 0.94 | 0.01 | 0.00 | 0.93 | 0.95 |
| Gemini-2.5-pro | 1 | 0.97 | 0.00 | 0.00 | 0.97 | 0.97 |
| Gemini-2.5-pro | 10 | 0.96 | 0.00 | 0.00 | 0.95 | 0.96 |
| Gemini-2.5-pro | 25 | 0.96 | 0.00 | 0.00 | 0.96 | 0.96 |
| Gemini-2.5-pro | 50 | 0.95 | 0.01 | 0.00 | 0.95 | 0.96 |
| Gemini-2.5-pro | 100 | 0.96 | 0.01 | 0.00 | 0.95 | 0.96 |
| Gemini-2.5-pro | 150 | 0.96 | 0.01 | 0.00 | 0.96 | 0.97 |
| Gemini-2.5-pro | 200 | 0.95 | 0.01 | 0.00 | 0.95 | 0.96 |
| Gemini-2.5-pro | 400 | 0.94 | 0.01 | 0.00 | 0.93 | 0.95 |
| Gemini-2.5-pro | 790 | 0.92 | 0.02 | 0.01 | 0.91 | 0.93 |
| GPT-5 mini (with thinking) | 1 | 0.98 | 0.00 | 0.00 | 0.98 | 0.98 |
| GPT-5 mini (with thinking) | 10 | 0.98 | 0.00 | 0.00 | 0.97 | 0.98 |
| GPT-5 mini (with thinking) | 25 | 0.97 | 0.00 | 0.00 | 0.96 | 0.97 |
| GPT-5 mini (with thinking) | 50 | 0.96 | 0.00 | 0.00 | 0.96 | 0.97 |
| GPT-5 mini (with thinking) | 100 | 0.96 | 0.01 | 0.00 | 0.96 | 0.96 |
| GPT-5 mini (with thinking) | 150 | 0.97 | 0.01 | 0.00 | 0.96 | 0.97 |
| GPT-5 mini (with thinking) | 200 | 0.96 | 0.01 | 0.00 | 0.95 | 0.96 |
| GPT-5 mini (with thinking) | 400 | 0.87 | 0.12 | 0.04 | 0.78 | 0.96 |
| GPT-5 (with thinking) | 1 | 0.98 | 0.00 | 0.00 | 0.98 | 0.98 |
| GPT-5 (with thinking) | 10 | 0.98 | 0.00 | 0.00 | 0.98 | 0.99 |
| GPT-5 (with thinking) | 25 | 0.98 | 0.00 | 0.00 | 0.98 | 0.98 |
| GPT-5 (with thinking) | 50 | 0.98 | 0.00 | 0.00 | 0.98 | 0.98 |
| GPT-5 (with thinking) | 100 | 0.98 | 0.00 | 0.00 | 0.98 | 0.98 |
| GPT-5 (with thinking) | 150 | 0.98 | 0.00 | 0.00 | 0.98 | 0.98 |
| GPT-5 (with thinking) | 200 | 0.98 | 0.00 | 0.00 | 0.98 | 0.99 |

### Geometric Mean √(Sensitivity × Specificity)

Geometric Mean $\sqrt{(\text{Sensitivity} \times \text{Specificity})}$
| AI Model | Batch Size | Mean | Std Dev | Std Error | Lower 95% CI | Upper 95% CI |
| --- | --- | --- | --- | --- | --- | --- |
| Gemini-2.5-flash | 1 | 0.91 | 0.00 | 0.00 | 0.91 | 0.91 |
| Gemini-2.5-flash | 10 | 0.95 | 0.01 | 0.00 | 0.94 | 0.95 |
| Gemini-2.5-flash | 25 | 0.94 | 0.01 | 0.00 | 0.94 | 0.95 |
| Gemini-2.5-flash | 50 | 0.95 | 0.01 | 0.00 | 0.94 | 0.95 |
| Gemini-2.5-flash | 100 | 0.94 | 0.02 | 0.01 | 0.92 | 0.96 |
| Gemini-2.5-flash | 150 | 0.95 | 0.02 | 0.01 | 0.94 | 0.96 |
| Gemini-2.5-flash | 200 | 0.95 | 0.01 | 0.00 | 0.94 | 0.96 |
| Gemini-2.5-flash | 400 | 0.96 | 0.01 | 0.00 | 0.95 | 0.96 |
| Gemini-2.5-pro | 1 | 0.98 | 0.00 | 0.00 | 0.98 | 0.98 |
| Gemini-2.5-pro | 10 | 0.97 | 0.00 | 0.00 | 0.97 | 0.98 |
| Gemini-2.5-pro | 25 | 0.98 | 0.00 | 0.00 | 0.97 | 0.98 |
| Gemini-2.5-pro | 50 | 0.97 | 0.00 | 0.00 | 0.97 | 0.98 |
| Gemini-2.5-pro | 100 | 0.98 | 0.00 | 0.00 | 0.97 | 0.98 |
| Gemini-2.5-pro | 150 | 0.98 | 0.00 | 0.00 | 0.98 | 0.98 |
| Gemini-2.5-pro | 200 | 0.97 | 0.01 | 0.00 | 0.97 | 0.98 |
| Gemini-2.5-pro | 400 | 0.96 | 0.01 | 0.00 | 0.96 | 0.97 |
| Gemini-2.5-pro | 790 | 0.94 | 0.01 | 0.00 | 0.94 | 0.95 |
| GPT-5 mini (with thinking) | 1 | 0.98 | 0.01 | 0.00 | 0.97 | 0.98 |
| GPT-5 mini (with thinking) | 10 | 0.98 | 0.00 | 0.00 | 0.98 | 0.99 |
| GPT-5 mini (with thinking) | 25 | 0.98 | 0.00 | 0.00 | 0.98 | 0.98 |
| GPT-5 mini (with thinking) | 50 | 0.98 | 0.00 | 0.00 | 0.97 | 0.98 |
| GPT-5 mini (with thinking) | 100 | 0.97 | 0.00 | 0.00 | 0.97 | 0.97 |
| GPT-5 mini (with thinking) | 150 | 0.97 | 0.01 | 0.00 | 0.96 | 0.98 |
| GPT-5 mini (with thinking) | 200 | 0.92 | 0.05 | 0.02 | 0.88 | 0.96 |
| GPT-5 mini (with thinking) | 400 | 0.65 | 0.04 | 0.01 | 0.62 | 0.68 |
| GPT-5 (with thinking) | 1 | 0.97 | 0.01 | 0.00 | 0.96 | 0.97 |
| GPT-5 (with thinking) | 10 | 0.98 | 0.01 | 0.00 | 0.98 | 0.99 |
| GPT-5 (with thinking) | 25 | 0.98 | 0.01 | 0.00 | 0.98 | 0.99 |
| GPT-5 (with thinking) | 50 | 0.98 | 0.00 | 0.00 | 0.98 | 0.99 |
| GPT-5 (with thinking) | 100 | 0.98 | 0.01 | 0.00 | 0.97 | 0.98 |
| GPT-5 (with thinking) | 150 | 0.98 | 0.01 | 0.00 | 0.97 | 0.98 |

### Gemini 2.5 Flash Detailed Results

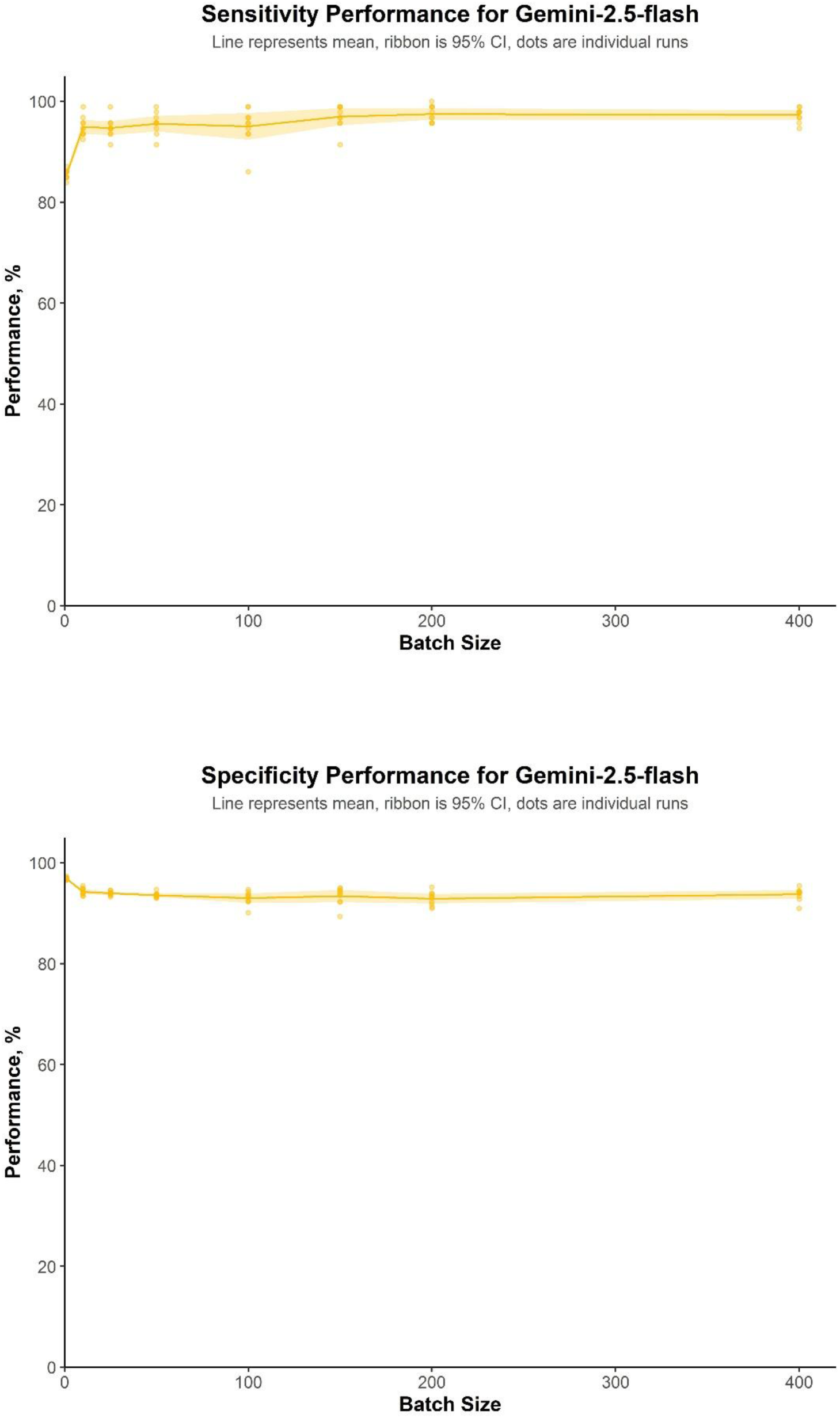

### 2.5 Pro Detailed Results

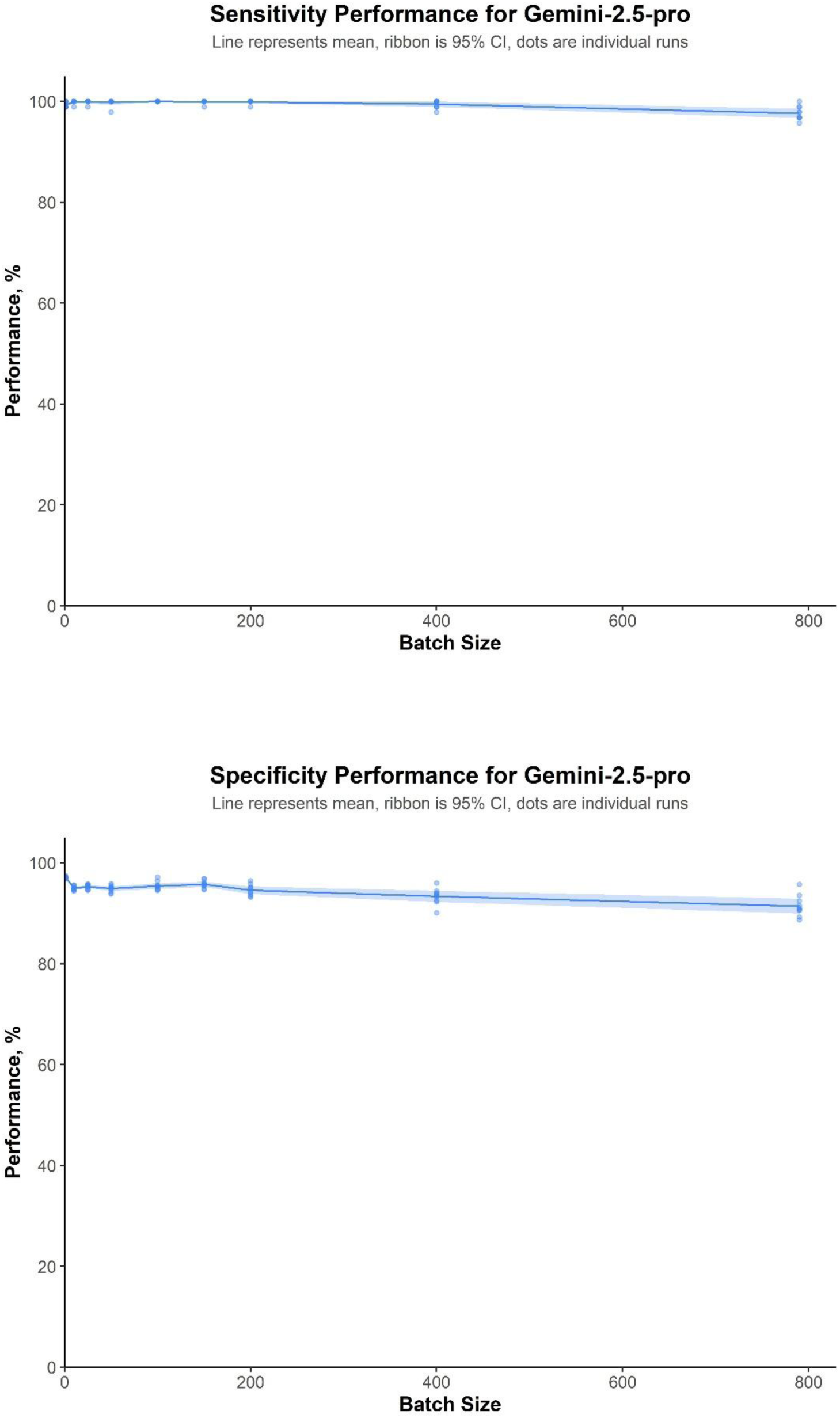

### GPT-5 (with thinking) Detailed Results

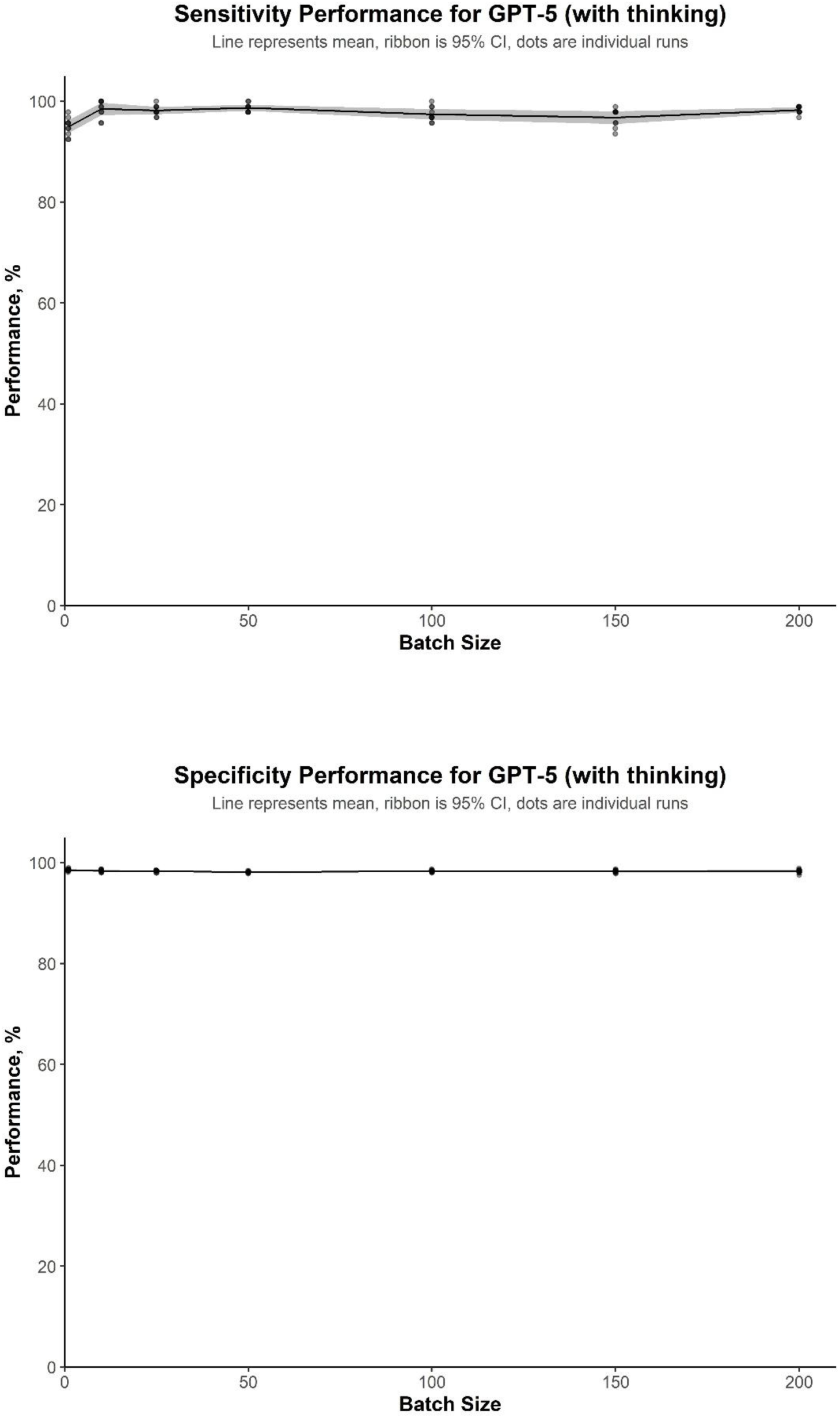

### GPT-5 mini (with thinking) Detailed Results

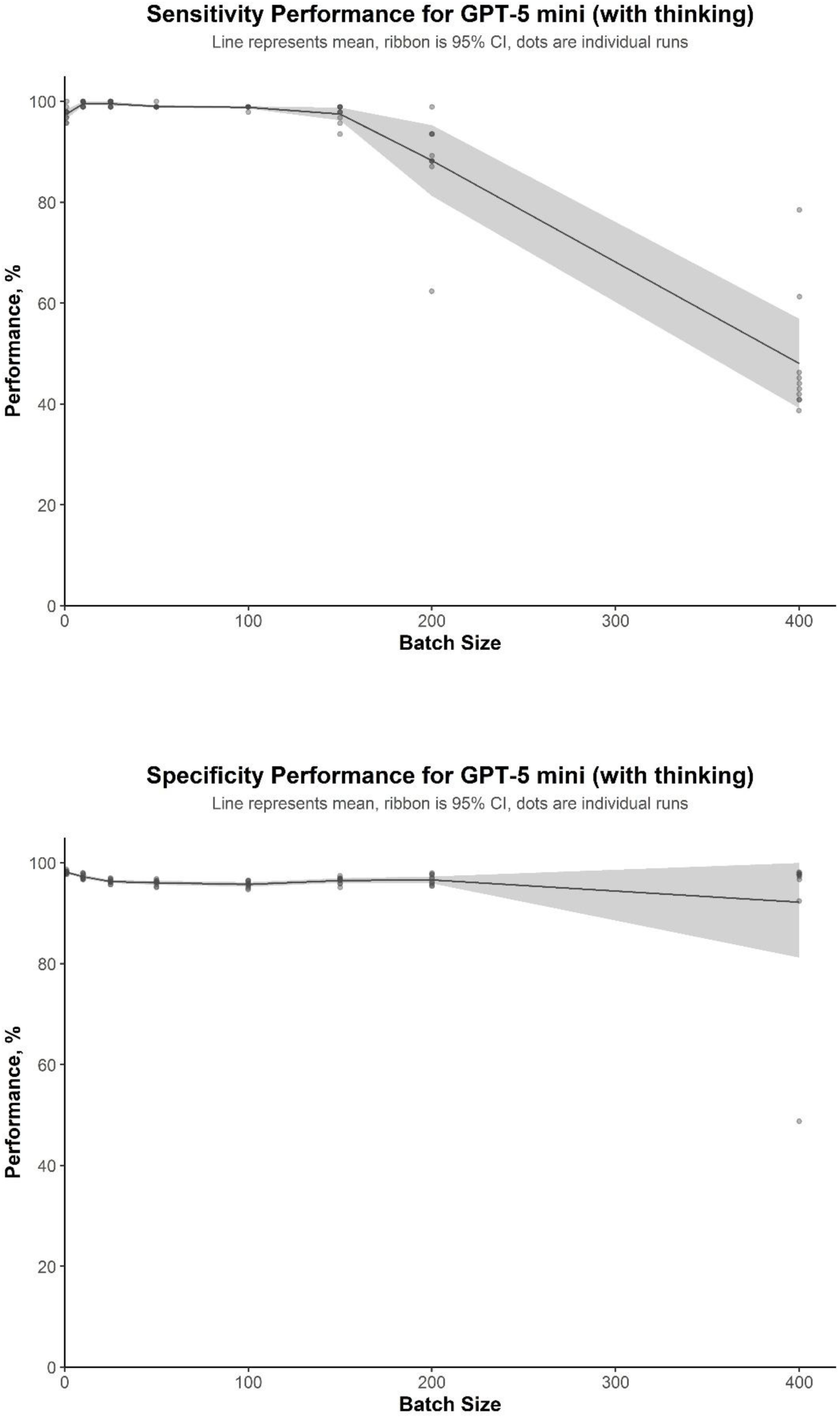

### Sensitivity and specificity for all tested models, including legacy models from OpenAI (o3 and GPT-4.1)

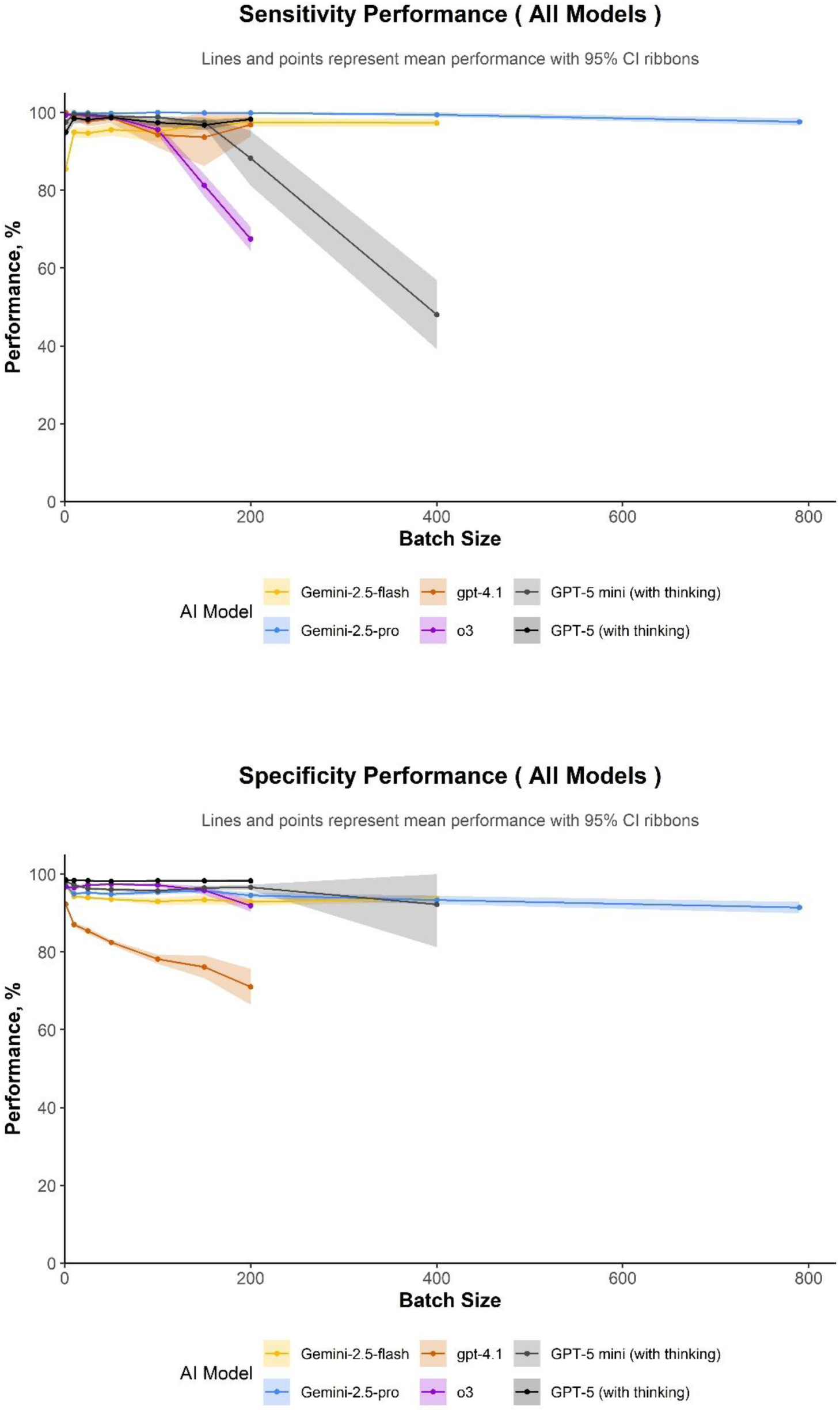

### Adjudication of citations initially classified as ’Include’ by the Cochrane authors

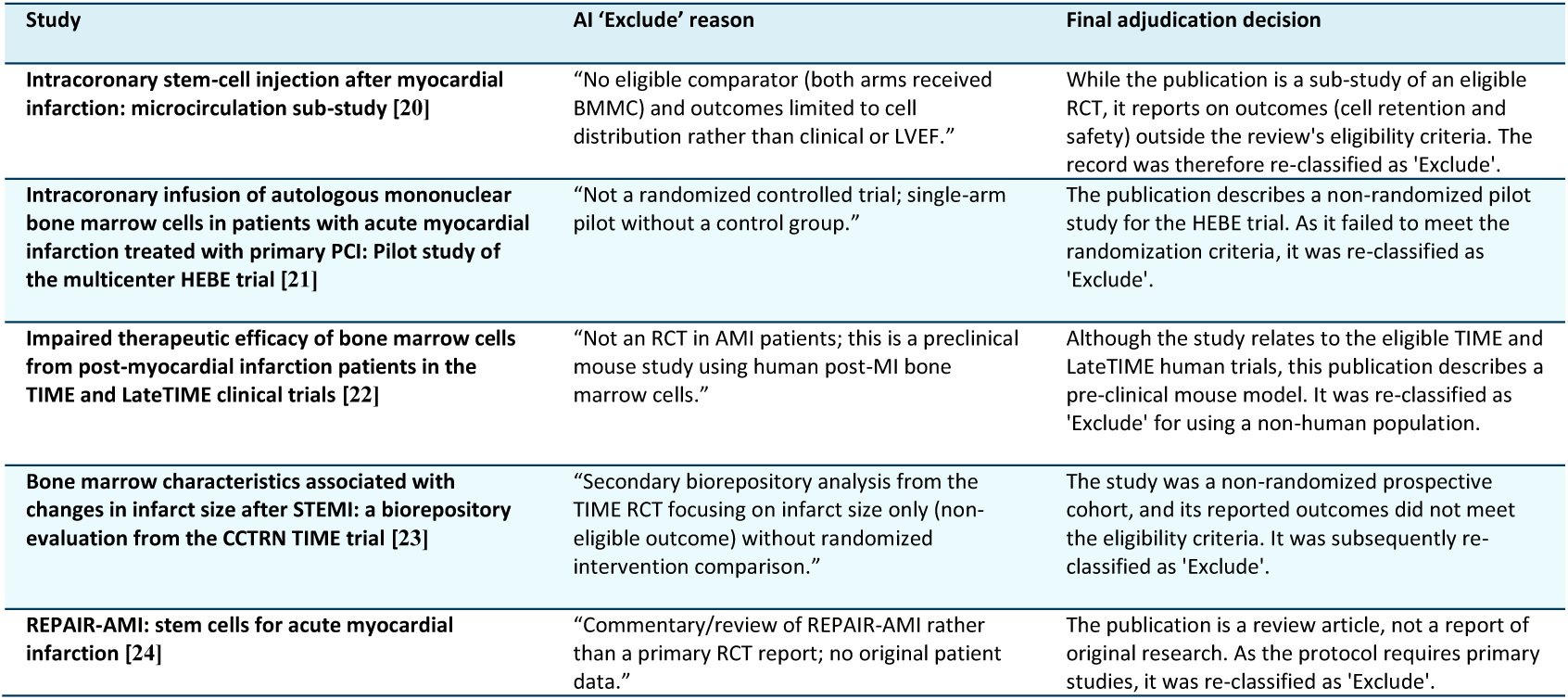

Adjudication of citations initially classified as 'Include' by the Cochrane authors
| Study | AI 'Exclude' reason | Final adjudication decision |
| --- | --- | --- |
| Intracoronary stem-cell injection after myocardial infarction: microcirculation sub-study [20] | "No eligible comparator (both arms received BMNC) and outcomes limited to cell distribution rather than clinical or LVEF." | While the publication is a sub-study of an eligible RCT, it reports on outcomes (cell retention and safety) outside the review's eligibility criteria. The record was therefore re-classified as 'Exclude'. |
| Intracoronary infusion of autologous mononuclear bone marrow cells in patients with acute myocardial infarction treated with primary PCI: Pilot study of the multicenter HEBE trial [21] | "Not a randomized controlled trial; single-arm pilot without a control group." | The publication describes a non-randomized pilot study for the HEBE trial. As it failed to meet the randomization criteria, it was re-classified as 'Exclude'. |
| Impaired therapeutic efficacy of bone marrow cells from post-myocardial infarction patients in the TIME and LateTIME clinical trials [22] | "Not an RCT in AMI patients; this is a preclinical mouse study using human post-MI bone marrow cells." | Although the study relates to the eligible TIME and LateTIME human trials, this publication describes a pre-clinical mouse model. It was re-classified as 'Exclude' for using a non-human population. |
| Bone marrow characteristics associated with changes in infarct size after STEMI: a biorepository evaluation from the CCTRN TIME trial [23] | "Secondary biorepository analysis from the TIME RCT focusing on infarct size only (non-eligible outcome) without randomized intervention comparison." | The study was a non-randomized prospective cohort, and its reported outcomes did not meet the eligibility criteria. It was subsequently re-classified as 'Exclude'. |
| REPAIR-AMI: stem cells for acute myocardial infarction [24] | "Commentary/review of REPAIR-AMI rather than a primary RCT report; no original patient data." | The publication is a review article, not a report of original research. As the protocol requires primary studies, it was re-classified as 'Exclude'. |

## Notes

### Competing Interest Statement

The authors have declared no competing interest.

### Funding Statement

This study did not receive any funding

